# Point of emission air filtration enhances protection of health care workers against skin contamination with virus aerosol

**DOI:** 10.1101/2021.08.15.21261997

**Authors:** Shane A Landry, Dinesh Subedi, Martin I MacDonald, Samantha Dix, Donna M Kutey, Jeremy J Barr, Darren Mansfield, Garun S Hamilton, Bradley A. Edwards, Simon A Joosten

## Abstract

**Rationale:** We recently demonstrated that a patient hood with a high efficiency particulate air filter eliminates virus aerosol contamination when very large quantities of bacteriophage virus are aerosolised into a clinical room. While this containment method is relatively low cost, it is unclear whether similar efficacy can be achieved with lower cost/commercial grade air purifiers, or if such an approach protects healthcare workers against virus aerosol contamination.

**Method:** A total of 10^9^ (10 ml of 10^8^) PhiX174 bacteriophages was nebulized into a sealed clinical room. Surface contamination was detected by settle plates left uncovered during exposure. A healthcare worker remained in the room, personal exposure was determined by skin swabs after exiting the room, following doffing of personal protective equipment (PPE). Four skin areas were swabbed: forearms/hands, neck, forehead, under N95 mask. Three conditions were tested, 1) hood with hospital grade air purifier (IQ Air Health Pro 250), 2) hood with commercial air purifier (Philips 1000i), and 3) control (no hood/air-purification).

**Findings:** The control condition demonstrated extensive environmental and limited skin contamination underneath PPE, which was highest under an N95 mask. The commercial air purifier and hood provided environmental control of virus aerosol and almost zero skin contamination. In comparison, the hospital grade purifier provided complete environmental and skin contamination protection, despite a lower clean air filtration rate (240m^3^/hr vs 270m^3^/hr). Virus counts on plates and swabs were significantly lower for both air purifiers and across neck, forehead, and under the N95. There were no statistically significant differences in detected virus counts between air purifiers.

**Conclusion:** This cheap and scalable method may be an effective way to reduce the spread of COVID-19 in hospitals by enhancing the effectiveness of PPE worn by health care workers who care for COVID-19 patients and who are exposed to virus aerosol.

Take home message
Relatively cheap portable air purifiers combined with a hood dramatically reduce the spread of virus aerosol and protect against environmental and healthcare worker contamination.

Plain Language Summary
This study shows that commercially available air purifiers, when combined with a hood that covers the head of a clinical bed, effectively capture very large amounts of virus aerosol in a simulated hospital setting. This virus containment strategy strongly reduced the number of viruses landing on surfaces in a clinical room. Crucially, this strategy also reduced that amount of virus detected on a healthcare worker’s skin underneath personal protective equipment, including under an N95 respirator. This cheap and scalable method may be an effective way to reduce the spread of COVID-19 in hospitals by enhancing the effectiveness of personal protective equipment worn by health care workers who care for COVID-19 patients and who are exposed to virus aerosol.

---

The World Health Organization (WHO) and Centres for Disease Control and Prevention (CDC) recently updated their advice regarding airborne transmission of SARS-CoV-2^1,2^, highlighting that virus laden aerosols can travel large distances and remain suspended in air for prolonged periods of time. Coupled with recent data suggesting that the Delta Variant of Concern is more transmissible^3^ and results in higher likelihood of admission to hospital^4^, the need to address virus aerosol transmission has never been greater.

Several studies have highlighted the effectiveness of aerosol control measures using point of emission air exchange/filtration. This strategy employs a containment structure (e.g. hood) and an expensive/hospital grade air purifier with a high efficiency particulate air (HEPA) filter. We recently demonstrated this method eliminates environmental contamination when very large quantities of bacteriophage virus are experimentally aerosolised into a non-ventilated clinical room^5^. While this method is relatively low cost compared to building/infrastructure alteration, it is unclear whether similar efficacy can be achieved with an ‘off the shelf’ air purifiers. Furthermore, it is not known if currently deployed personal protective equipment (PPE) strategies protect against virus aerosol transmission to healthcare workers, or if point of emission control of virus aerosol can enhance the effectiveness of PPE.

In this context we used a bacteriophage “live” virus model of aerosol transmission to:

1. Assess the ability of an “off the shelf” air purifier and hood to reduce environmental contamination,
2. Assess the effectiveness of a commonly deployed PPE strategy to protect against skin contamination,
3. Determine if - the protection offered by PPE can be enhanced by a point of emission aerosol control strategy.

Utilizing our previously described method^5^ we systematically tested virus aerosol surface contamination of a clinical room and skin contamination of a healthcare worker wearing a gown (Jiangxi Fashionwind Apparel Co. Ltd.), disposable gloves (Mediflex Industries), face shield (Xamen Sanmiss Bags Co.) and an N95 respirator (BYD Precision Manufacture Co., Ltd.). We nebulised (Pari-Pep S System, PARI) a total of 10^9^ (10 ml of 10^8^) PhiX174 bacteriophages into a sealed clinical room with dimensions: 4.0 × 3.25 × 2.7 m (surface area=13.0 m^2^, volume=35.1 m^3^). Surface contamination was detected by 13 soft agar overlays containing *Escherichia coli C* bacterial host left uncovered for the duration that bacteriophage lysate was nebulized (∼40 mins). Plates were sealed after nebulisation and new plates were exposed over two consecutive 15 min intervals after nebulisation to quantify residual virus settling. After a total exposure period of 70 mins the healthcare worker exited the room. Personal contamination was determined by skin swab following doffing of PPE. The doffing procedure was video recorded and examined independently by two expert nurses to ensure doffing procedure compliance. Doffing occurred in a clinical room separated from the testing room by a corridor and 4 sealed doors. The doffing room had continuous HEPA filtration (5 exchanges per hour) applied at all times. Control plates were opened at the time of doffing and swabbing to determine if any viruses were present/settling during the doffing/swabbing procedure. Swabs (Jumbo Swabs, Multigate Medical Products Pty Ltd.) were individually immersed in 3 mL of 1X phosphate buffered saline (PBS) contained in a test tube, and then applied individually and systematically to four separate areas – 1) forearms and back of hands, 2) neck, 3) forehead, and 4) under the N95 mask (around mouth/nose under mask coverage). Swabs were re-immersed in the PBS within the test tube, vigorously mixed with 1 mL of PBS collected and plated neat with bacterial host to obtain virus count from each individual swab.

Surface and personal contamination was assessed across 3 experimental conditions:

1. No air filtration or hood structure applied (control condition).
2. Air filtration and hood applied with “off the shelf” HEPA filter applied at 270m^3^/hr (Philips, Amsterdam, Netherlands).
3. Air filtration and hood applied with hospital grade HEPA filter set to 240m3/hr (Swiss Made, Goldach, Switzerland).

Each experimental condition was repeated 3 times. Kruskal-Wallis with uncorrected-Dunn’s post-hoc test was used to compare virus counts between conditions for settle plates and for each skin swab area. The room was purged for 30mins using the hospital grade HEPA filter set to 470m^3^/hr. Control plates were left uncovered post-purge to confirm decontamination was complete.

The control condition demonstrated extensive environmental and also limited skin contamination underneath PPE, which was highest on the nose/mouth (Figure 1). The “off the shelf” air purifier filter and hood provided environmental control of virus aerosol and almost zero skin contamination. In comparison, the hospital grade air purifier provided complete environmental and skin contamination protection, despite a lower clean air filtration rate (240m^3^/hr vs 270m^3^/hr). Virus counts on plates were significantly lower for both air purifiers across all three time intervals. Similarly, virus counts from skin swabs were significantly lower on neck, forehead and under the N95 (all p<0.05). There we no statistically significant differences in detected virus counts between the IQAir and Philips 1000i air purifiers.

**Figure 1.**
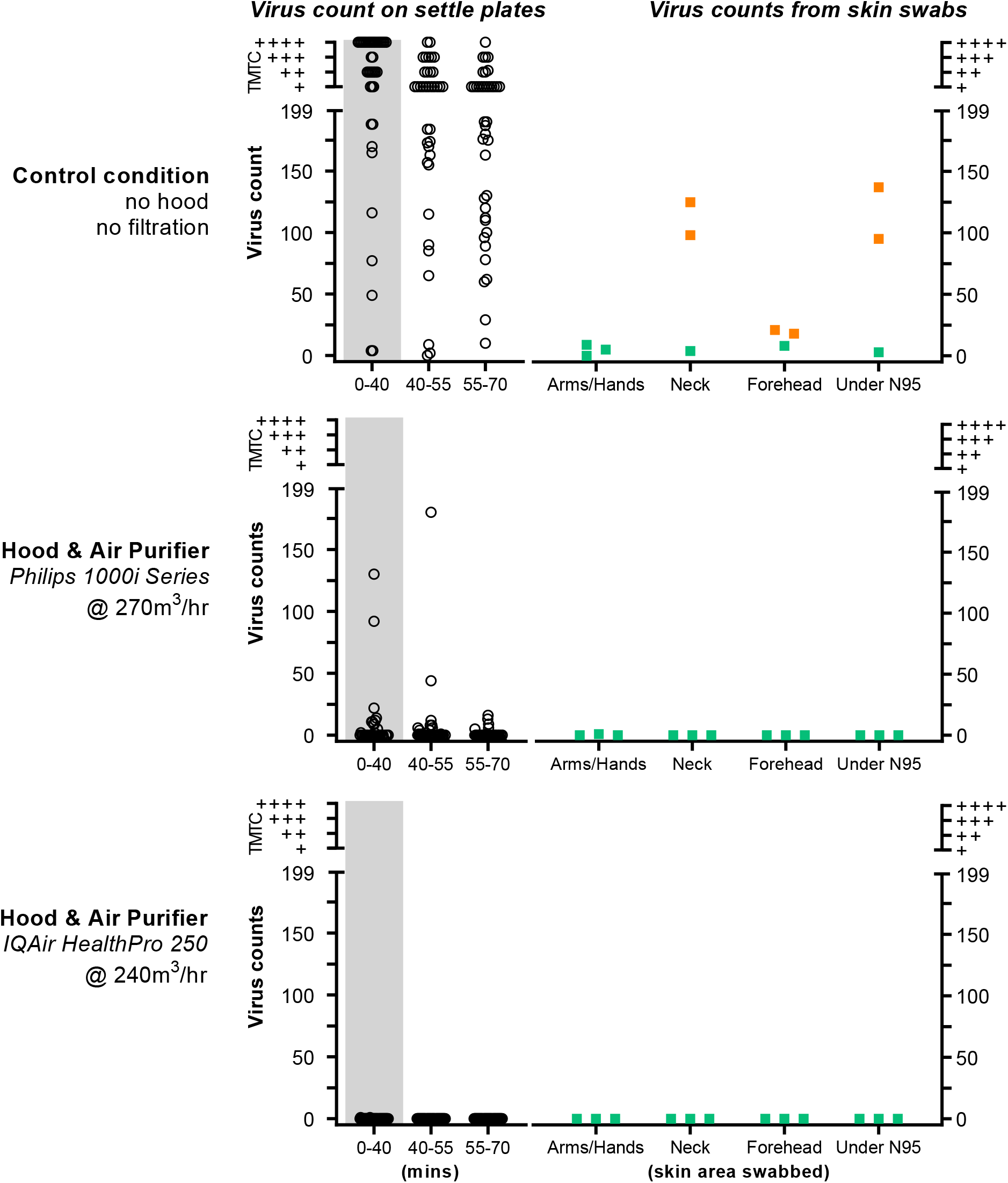
Virus plaque counts per experimental condition. Graphs on the left quantify environmental contamination in the clinical room from virus aerosol. Open circles represent virus counts on settling plates, closed circles show plates within 1m of the aerosol source. Grey bars represent the period of active nebulisation (40mins). Plates were closed and new ones reopened over two 15min intervals after nebulisation to quantify residual virus settling over time. Virus counts were quantified as plaque forming units (PFU) as previously described^9^. Virus counts >200 were considered too-many-to-count (TMTC) and were rated using an ordinal (+, ++, +++, ++++) visual rating scale. Squares on the right show virus counts determined from skin surface swabs for each condition. Squares are also coded green and amber to reflect qualitative ratings of mild (≤10) and intermediate (11-199) virus counts.

To our knowledge this is the first study to explore the interaction between air purification and PPE in protecting against virus aerosol. We demonstrate that widely used PPE provides incomplete protection against skin contamination from prolonged (70 mins) exposure to an environment with a high number of virus laden aerosols and poor ventilation. Moreover, skin contamination was greatest on the face, beneath a non-fit-tested N95 respirator. This elevated level of contamination compared to other skin sites is likely due to the suction produced by the healthcare worker’s respiration (which does not affect other sites). These data demonstrate that the effectiveness of PPE in preventing skin contamination is enhanced by the use of a hood and HEPA filter point of emission control strategy. Even an “off the shelf” HEPA filter demonstrated a large impact on virus aerosol contamination.

The COVID-19 pandemic has led to a complete re-appraisal of the science and assumptions underpinning infection control practice and virus aerosol transmission. The CDC and WHO now recognise the key importance of virus aerosol transmission to the spread of SARS-CoV-2^1,2^. Critical to the impact of this mode of transmission is emerging evidence that patients who acquire SARS-CoV-2 infections in hospital have a mortality rate of approximately 30%^6,7^. Furthermore, thousands of healthcare workers who have been infected with SARS-CoV-2 while treating patients in their workplace have subsequently died from COVID-19. The need to address airborne virus spread in our hospitals is critical. Our data provide evidence that a simple, cost effective, and scalable approach utilizing a containment at point of emission strategy can negate environmental contamination by virus aerosol and can enhance the effectiveness of PPE in protecting against skin contamination.

Our study has some important limitations. Firstly, we deployed a single approach to PPE that utilized a non-fit-tested N95 respirator. Future work is needed to systematically assess the effectiveness of various mask strategies (e.g. fit-tested N95 respirators) in preventing skin contamination of the face. Secondly, we believe that skin contamination detected underneath the N95 respirator most likely represents the “tip of the iceberg” of likely virus aerosol contamination of the airway. Given that the nebulizer we use produces 3.4µm particles^8^, it is likely many of these are being inhaled into the upper and lower respiratory tract beneath the N95 respirator. Both of these issues are ongoing areas of research by our group.

## Data Availability

Data will be available on reasonable request.

